# What are the most effective interventions to support children and young people bereaved by suicide in the family: a rapid review

**DOI:** 10.1101/2023.09.13.23295481

**Authors:** Mala Mann, Meg Kiseleva, Lydia Searchfield, Francesca Mazzaschi, Rhiannon Jones, Kate Lifford, Alison Weightman, Ann John, Ruth Lewis, Rhiannon Tudor Edwards, Jacob Davies, Alison Cooper, Adrian Edwards

**Affiliations:** Specialist Unit for Review Evidence, Cardiff University, United Kingdom; Health and Care Research Wales Evidence Centre, Cardiff University, United Kingdom; School of Health and Social Care, Swansea University Medical School, United Kingdom; Health and Care Research Wales Evidence Centre, Bangor University, United Kingdom

## Abstract

Bereavement by suicide is different from other forms of bereavement and needs specialised support. Children and young people who lost loved ones to suicide are more likely to suffer a complicated bereavement process and have poorer mental health.

This review aims to assess the evidence for the effectiveness of interventions to support children and young people (up to the age of 24 years) bereaved by suicide. The review included evidence available up until 29 March 2023. Three studies were identified and all reported on group therapy interventions lasting between 10 and 14 weeks.

**Key findings and certainty of the evidence:** Reductions in anxiety and depressive symptoms were found in children who received the group interventions. However, due to the types of study designs used and limitations of the included studies, it is unclear if this is attributable to the interventions, so caution should be applied when generalising the results.

The strongest evidence came from a non-randomised controlled study, in which children in the intervention group had significantly greater reduction of anxiety and depressive symptoms compared with children in the control group. However, this study was limited due to numbers of participants lost to follow-up.

**Research Implications and Evidence Gaps:** Further research is needed to develop interventions to support children and young people bereaved through death by suicide of a family member. Additional research is needed to evaluate the effectiveness and cost-effectiveness of planned interventions.

**Policy and Practice Implications:** It is difficult to draw firm conclusions due to the limited evidence and low quality of included studies. However, there are indications that group interventions may help to reduce anxiety and depressive symptoms in children bereaved by suicide. It will be important to develop guidance and standards of practice for these services based on best available evidence. All such services must use validated outcome measures as part of an integral evaluation process set up from service initiation.

**Funding statement:** The Specialist Unit for Review Evidence was funded for this work by the Health and Care Research Wales Evidence Centre, itself funded by Health and Care Research Wales on behalf of Welsh Government

## 1. BACKGROUND

### 1.1 Who is this review for?

This Rapid Review was conducted as part of the Health and Care Research Wales Evidence Centre Work Programme. The question for this review was suggested by the Suicide and Self-Harm Prevention team in the NHS Wales Executive to support the Welsh Government’s suicide prevention strategy objectives to provide information and support to those bereaved or affected by suicide and self-harm; to build on the recent consultation on draft guidance on how we respond to people affected or bereaved by suicide; and to inform the development of the new Suicide and Self-harm Prevention Strategy. The findings will guide the Government’s work with agencies and charities that support children and young people following the suicide of a close family member.

### 1.2 Background and purpose of this review

In 2021, there were 5,583 suicide deaths in England and Wales (Office for National Statistics, 2022). Many of these deaths would have been parents or siblings, leaving behind bereaved children and young people.

Bereavement by suicide is different from other forms of bereavement, and the need for specialised support is indicated (Braiden et al, 2009; Pfeffer et al, 2002). Children who lost loved ones to suicide are more likely to experience a complicated or complex bereavement process and have poorer mental health outcomes (Andriessen et al, 2016; Braiden et al, 2009; Pitman et al, 2014). Compared to others, children and young people bereaved by family suicide are at higher risk of dying by suicide (Calderaro et al, 2021; Del Carpio et al, 2021; Hua et al. 2019) and experiencing a range of other negative outcomes, such as attempting suicide (Calderaro et al, 2021; Del Carpio et al, 2021) and self-harm (Del Carpio et al, 2021).

Prior to this review, an initial search for secondary evidence (systematic reviews, rapid reviews and scoping reviews) revealed a number of relevant reviews (Andriessen et al, 2019; Bergman et al, 2017; Chen and Panebianco, 2018; Hua et al, 2020; Journot-Reverbel et al; 2017, Kaspersen et al, 2022; Ridley and Frache, 2020). However, only one review (Journot-Reverbel et al. 2017) was specific to the inclusion criteria. The other reviews included adults as well as children and adolescents, or children and adolescents bereaved by causes other than suicide. The primary studies reported in the Journot-Reverbel et al. (2017) review (Daigle and Labelle, 2012; Pfeffer et al, 2002) were included in this work, as well as an additional study. The search conducted for this review also covered a more recent period up to March 2023.

## 2. RESULTS

### 2.1 Included studies

A search for primary studies identified 348 records, of which 3 studies met the inclusion criteria (Section 5.1) and were included in this review. Those were two studies with a pre-post design and one non-randomised controlled study. See Section 6.2 for a detailed summary of the included studies.

#### 2.1.1. Study description

The non-randomised controlled trial (Pfeffer et al, 2002) was based in the USA. This study reported on an intervention that included 10 weekly 1.5-hour group sessions which involved psychoeducational and supportive components. The aim of the intervention was to promote children’s healthy adjustment after family suicide and to reduce morbid outcomes. Semi-structured interviews were conducted pre– and post-intervention to measure psychosocial variables with time between interviews being between 2.5-4.5 months. The study included 75 children initially (39 children in the intervention group and 36 children in the control group), but saw a high drop-out rate, with 32 children retained in the intervention group and 9 children in the control group. The children were aged between 6 and 15 years old and split into three age groups for the intervention.

One uncontrolled pre-post study (Daigle and Labelle, 2012) reported a pilot evaluation of the Group Therapy Program for Children Bereaved by Suicide based in Canada. This intervention includes 12 two-hour group therapy sessions over 14 weeks which aim to help children and their surviving parents cope with the difficulties of the grieving process. This was evaluated using observation checklists during the programme as well as psychological and social assessments two to three weeks before and one to two weeks after. The study included 8 children aged 6 to 12 years old.

The other uncontrolled pre-post study (Veale et al, 2014) was based in Ireland. This was a group intervention comprising of therapeutic groupwork of 1.5 hours per week over 12 weeks including art, physical, reflective and mindfulness activities. This intervention aimed to progressively explore the bereavement experience, moving to memories of the loved ones and finally a focus on the future. Behavioural, psychological and social outcomes were assessed pre-intervention, post-intervention, and then during six-month and four-year follow-ups.

The studies are summarised in Table 1.

**Table 1.**
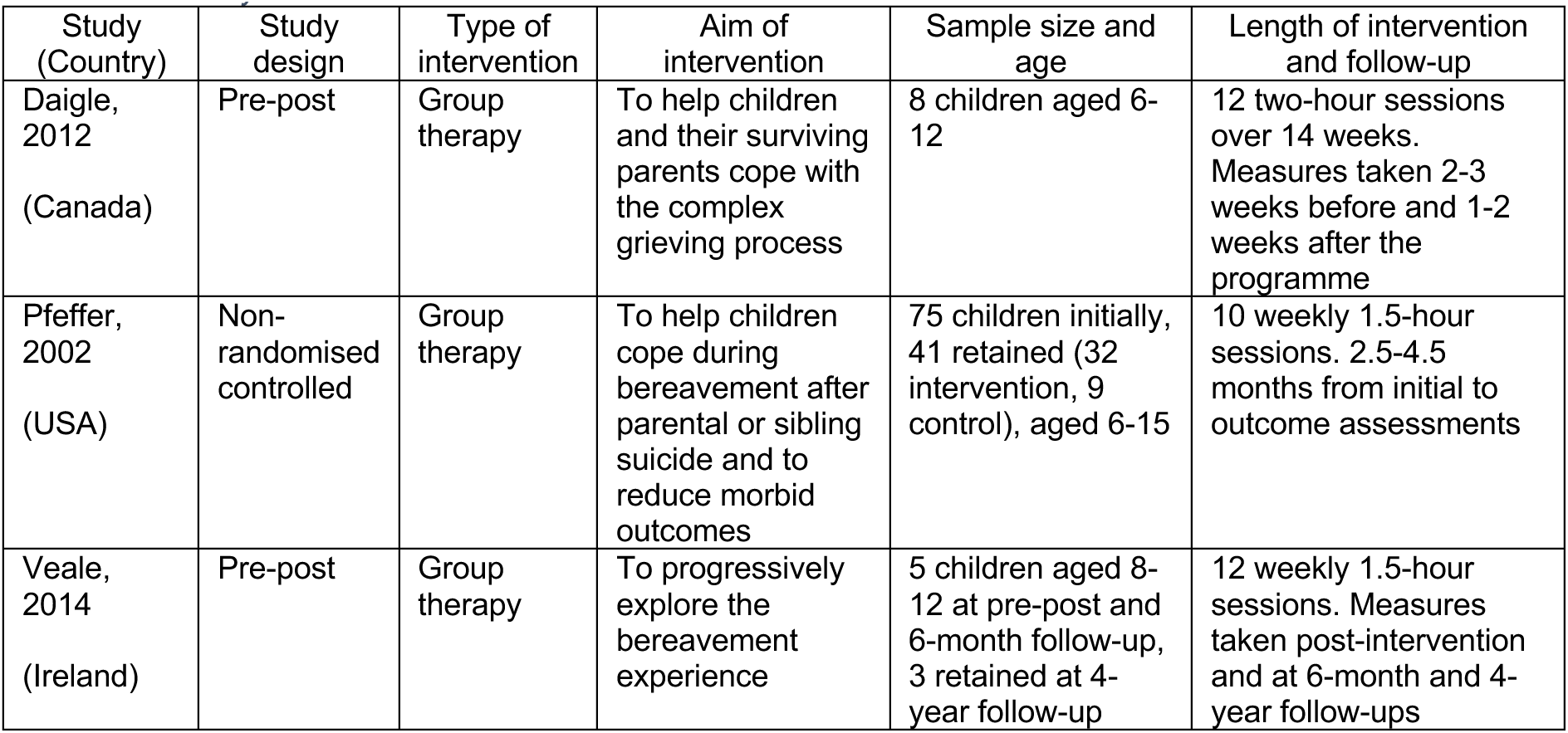
Summary of interventions.

#### 2.1.2. Outcomes measured

These studies covered a variety of mental health, social and behavioural outcomes. The number of studies that measured each outcome is summarised in Table 2.

**Table 2.** Summary of outcomes reported in included studies.

| Category | Outcome | No of studies |
| --- | --- | --- |
| Mental Health outcomes | Anxiety | 2<sup>1,2</sup> |
|  | Depression | 2<sup>1,2</sup> |
|  | Internalising | 1<sup>3</sup> |
|  | General mood | 1<sup>1</sup> |
|  | Intrapersonal EI | 1<sup>1</sup> |
|  | Posttraumatic stress | 1<sup>2</sup> |
|  | Grief | 1<sup>1</sup> |
| Social and behavioural outcomes | Anger | 1<sup>1</sup> |
|  | Disruptive behaviour | 1<sup>1</sup> |
|  | Externalising | 1<sup>3</sup> |
|  | Stress management | 1<sup>1</sup> |
|  | Adaptability | 1<sup>1</sup> |
|  | Competence (activities, school, social) | 1<sup>3</sup> |
|  | Task difficulty | 1<sup>3</sup> |
|  | Hope | 1<sup>1</sup> |
|  | Self-esteem | 1<sup>1</sup> |
|  | Social adjustment | 1<sup>2</sup> |
|  | Relationships with parents | 1<sup>3</sup> |
|  | Interpersonal EI | 1<sup>1</sup> |

EI = Emotional intelligence. <sup>1</sup>Daigle (2012), <sup>2</sup>Pfeffer (2002), <sup>3</sup>Veale (2014).

### 2.2 Effectiveness of group interventions for mental health outcomes

All three studies included in this review measured children’s mental health outcomes, although specific outcomes varied by study. Anxiety and depression were measured in two studies (Daigle and Labelle, 2012; Pfeffer et al, 2002). Other mental health outcomes, such as internalising symptoms, general mood, posttraumatic stress, and grief, were each measured in one study.

#### 2.2.1. Anxiety, depression and general mood

Both studies that examined changes in anxiety (Daigle and Labelle, 2012; Pfeffer et al, 2002) observed improvements in participating children’s outcomes. There was a greater reduction (*p* ≤ .004) and rate of reduction (*p* ≤ .01) in the intervention group than in the control group in a controlled study of children aged 6–15 (Pfeffer *et al*, 2002). Significant reduction in anxiety was noted in the intervention group between the assessments before (T1) and after (T2) the intervention (*F*1,25 = 4.6, *p* ≤ .04). A pre-post study of children aged 6-12 also saw a 13.65% reduction in anxiety, but no statistical tests were conducted due to the small sample size (Daigle and Labelle, 2012).

Two studies measured depression scores pre– and post-intervention (Daigle and Labelle, 2012; Pfeffer et al, 2002). Both reduction of depression (*p* ≤ .0003) and the reduction rate (*p* ≤ .0006) were greater in the intervention group than in the control group and there was a significant decrease in depression from T1 to T2 (*F*1,26 = 10.6, *p* ≤ .003) in the intervention group in the controlled study (Pfeffer et al, 2002). Daigle and Labelle (2012) also noted a 11.99% reduction in depressive symptoms, but the small sample size precluded testing whether the difference was statistically significant.

In addition, the pre-post study conducted by Veale (2014) reported internalising scores in children aged 8–12. Out of the five children that participated, pre-intervention, four scored within the clinical range for internalising and one had a borderline score. Post-intervention, one child was in the clinical range and two had borderline scores, which remained the case at 6 months after the intervention. At the 4-year follow-up, in which three out of the five children took part, all three scored within the normal range.

Children’s general mood pre– and post-intervention was measured in one study, which saw a 3.08% improvement in their scores (Daigle and Labelle, 2012). At the same time, children’s intrapersonal emotional intelligence, i.e., their ability to understand their emotions, increased by 6.34%. No statistical tests were conducted.

#### 2.2.2. Posttraumatic stress and grief

Changes in posttraumatic stress were measured in one study, which observed no significant change within the intervention group and no significant differences in outcome scores or rates of change between the intervention and the control groups (Pfeffer et al, 2002). However, children’s grief symptoms were reduced by 29.52% for children taking part in another group intervention (Daigle and Labelle, 2012).

#### 2.2.3. Bottom line results for mental health outcomes

Overall, the three studies included in this review reported improvements in mental health outcomes, however, the methodological limitations of these studies, such as small sample sizes and the lack of a control group in the pre-post studies (Daigle and Labelle, 2012; Veale, 2014) as well as the lack of randomisation, the high drop-out rate, and the differences between the intervention and control groups in the controlled study (Pfeffer et al, 2002) limit the generalisability of the findings.

### 2.3 Effectiveness of group interventions for social and behavioural outcomes

All three studies examined at least one social or behavioural outcome of participating children, such as anger, disruptive behaviour, stress management, adaptability. Each of the outcomes reported below was only reported in one study.

#### 2.3.1. Anger and disruptive behaviour

One study examined changes in children’s anger and disruptive behaviour (Daigle and Labelle, 2012). They observed a 10.42% reduction in disruptive behaviour, but a 7.91% increase in anger scores. However, another study, also using a pre-post design, saw a general decrease in externalising symptoms, with two of the five participating children in the clinical range post-intervention and three children having borderline scores at 6 months, compared to three children in the clinical range before the intervention (Veale, 2014). At the 4-year follow-up, all three children who took part scored in the normal range.

#### 2.3.2. Stress management and adaptability

Stress management and adaptability were examined in one pre-post study (Daigle and Labelle, 2012). Improvements were observed in both outcomes (+4.24% and +4.45% respectively). However, no statistical tests were conducted due to sample size limitations.

#### 2.3.3. Competence and self-esteem

A pre-post study of five children measured their competence in performing activities, school tasks, and social functions (Veale, 2014). Only one of the five children scored in the clinical range pre-intervention and all five children had scores in the normal range post-intervention and at the 6-month follow-up. Three children participated in the 4-year follow-up and had scores in the normal range.

Children also received a functional assessment within the same intervention which measured their perceived levels of difficulty completing various tasks. Three out of the five children said that most or all tasks were easier six months post-intervention, but there was no clear pattern to these changes pre-intervention to immediately after the intervention, to six months later.

Children’s hope about achieving their goals was examined in one study (Daigle and Labelle, 2012). The questions related to their goal-directed determination and planning of ways to meet their goals. There was an improvement of 11.57% and 18.58% on these dimensions respectively, as well as an overall improvement of 14.85%. In addition, there was a 12.44% improvement in children’s self-esteem pre– and post-intervention.

#### 2.3.4. Social relationships

There were no differences in social adjustment before and after the intervention, or between the intervention and control groups at T2, for children participating in a controlled study of a group intervention (Pfeffer et al, 2002). However, in the other studies, improvements in relationships and interpersonal emotional intelligence were observed. In a pre-post study, four out of five participating children described their relationships with their parent(s) as good post-intervention and at the 6-month follow-up, compared to three children pre-intervention (Veale, 2014). Children’s interpersonal emotional intelligence saw a 9.71% increase in the other pre-post study (Daigle and Labelle, 2012).

#### 2.3.5. Bottom line results for social and behavioural outcomes

All outcomes reported in this section apart from social adjustment were examined in pre-post studies (Daigle and Labelle, 2012; Veale, 2014), so no control group was present. In addition, the sample sizes in these studies were severely limited, precluding the possibility of conducting statistical tests. This limits the generalisability of the findings.

## 3. DISCUSSION

### 3.1 Summary of the findings

Limited evidence is available on support interventions for children and young people bereaved by suicide. Only three comparative studies examining such interventions were located. Overall, the interventions reviewed in this report showed promising results, but the methodological limitations preclude any definitive conclusions. No evidence was found on support interventions for very young children below the age of 6 or young people older that 15, but below 24.

All three included studies (Daigle and Labelle, 2012; Pfeffer et al, 2002; Veale, 2014) reported on group interventions. The most common outcomes examined by the studies included in this review were related to the symptoms of anxiety and depression, with two studies (Daigle and Labelle, 2012; Pfeffer et al, 2002) examining changes specifically in anxiety and depression, and one more study (Veale, 2014) reporting on internalising symptoms. All three studies observed improvements, however, the absence of a control group in the pre-post studies makes it impossible to determine that the changes were attributable to the interventions. Other types of outcomes (such as anger and disruptive behaviour, self-esteem, social adjustment) were only examined in one study each.

Though the non-randomised controlled study (Pfeffer et al, 2002) provided stronger evidence for the efficacy of a group intervention for reducing anxiety and depressive symptoms, several methodological problems were identified, including alternating (rather than random) assignment, differences between groups, and major dropout among non-intervention families (75% vs 18% among families who received the intervention). The two pre-post studies (Daigle and Labelle, 2012; Veale, 2014) observed positive changes in the participating children in terms of change percentage and the number of children in the clinical range for various outcomes, however, the lack of statistical analysis due to the small sample size and the lack of a control group were key limitations.

While this review captured only those studies where the outcomes of interventions for bereaved children were measured quantitatively, it is helpful to turn to studies employing qualitative design to understand why such interventions may be effective. In addition to the content of the intervention, the context in which it takes place is important. For example, bereaved children who participated in group interventions stated that such programmes helped them to feel less alone as they had the opportunity to meet others who shared the same experience and could therefore understand what they were going through (Braiden et al, 2009; Hagstrom, 2021; McClatchey and Wimmer, 2012). Being able to meet others who had been bereaved by suicide and openly talk about a loved one’s death helped some participants destigmatise their understanding of suicide (Hagstrom, 2021). They were also able to see how other children and young people were dealing with the suicide (McClatchey and Wimmer, 2012). This suggest that the group aspect is an important part of the intervention. Even though these qualitative studies do not fit the eligibility criteria for this review, the summary of their results is provided in Appendix 3.

### 3.2 Strengths and limitations of the available evidence

The evidence presented in this review has a number of limitations. Two of the three included studies (Daigle and Labelle, 2012; Veale, 2014) used a pre-post design with no control group and had small sample sizes (8 and 5 children respectively), meaning that their findings may not be generalisable to other contexts. Only one controlled study of an intervention for suicide-bereaved children was found (Pfeffer et al, 2002), however, the children were not randomly assigned to the intervention and control groups and the study saw a large dropout rate, particularly in the control group. While all three studies observed improvements in children’s outcomes, their findings should be treated with caution due to the methodological limitations of these studies.

### 3.3 Strengths and limitations of this Rapid Review

Although this review was conducted rapidly to inform policy and decision makers, comprehensive search strategies were designed to identify relevant evidence in the bibliographic databases. Database searches were supplemented by searching a range of websites known to review team and the stakeholders as being potentially relevant.

In conducting this rapid review, two reviewers independently carried out study selection. However, quality appraisal and data extraction extraction were carried out by single reviewers and independently checked for accuracy. Any disagreements were resolved through discussion. The review consists of 3 primary studies conducted in Ireland, Canada and the USA. All included studies had significant limitations. The primary weakness is the sample size and lack of a control group. Due to the paucity of evidence, we were unable to undertake any assessment of the outcomes using GRADE.

This review highlights that there is a lack of research in this area and the need for further research into the development and evaluation of interventions.

### 3.4 Implications for policy and practice

Given the limited evidence available on support interventions for children and young people bereaved by suicide, it will be important to develop guidance and standards of practice for these services based on best available evidence. All services developed will need to have measurable validated outcomes as part of an integral evaluation process set up from service initiation.

These findings will be presented and discussed at the National Advisory Group to Welsh Government for Suicide and Self-Harm Prevention and will inform the development of guidance setting out a systems response to children and young people affected or bereaved by suicide.

### 3.5 Implications for future research

There is a need for high quality primary studies examining the effectiveness of bereavement support interventions for children and young people bereaved through the suicide of a family member. These studies should aim for rapid translation of evidence into practice to ensure service improvement and involvement of children and young people, highlighting important aspects of an intervention as well as its delivery.

### 3.6 Economic considerations*

- An estimated 300-350 people die by suicide in Wales each year (Samaritans 2017). Inflated to July 2023 prices, these suicides cost the Welsh economy between £760million and £880million per annum. Approximately 70% of these costs are attributed to emotional impacts on family and on society (Public Health England, 2020; Bank of England 2023).
- Suicide risks increase during period of economic recession. Economic downturns categorised by rising unemployment further exacerbate this risk. The lag in macroeconomic conditions improving after recession can lead to these suicide risks persisting for several years, especially if personal circumstances have not improved (Samaritans, 2017).
- The attempted suicide or loss of a parent to suicide are categorised as Adverse Childhood Experiences (ACEs). An evidence base exists around economic impacts of ACEs more generally (Hughes et al., 2021).
- Early parental death (before the age of 21) was found to be consistently associated with higher risk of hospitalisation and higher medication use for mental health disorders as well as increased work absenteeism due to illness in adulthood (Böckerman, Haapanen and Jepsen, 2023). These risk factors in turn can incur additional costs to society through additional resource use cost and productivity losses. Early parental death was also linked to a significant reduction in years of school attendance, employment and earnings in adulthood. Although the detrimental impacts were observed for both males and females, the negative consequences for parental death were greater for males (Böckerman et al., 2023).

\**This section has been completed by the Centre for Health Economics & Medicines Evaluation (CHEME), Bangor University*

## Data Availability

All data produced in the present study are available upon reasonable request to the authors

## Abbreviations

CI: Confidence Interval
EI: Emotional intelligence
p: Probability
M: Mean/Average
NR: Not reported
RCT: Randomised Controlled Trial
RR: Relative Risk
SD: Standard deviation
T1/T2: Time interval (pre-intervention (T1) / (post-intervention (T2)
USA: United States of America

## 5. RAPID REVIEW METHODS

### 5.1 Eligibility criteria

**Table 3.**
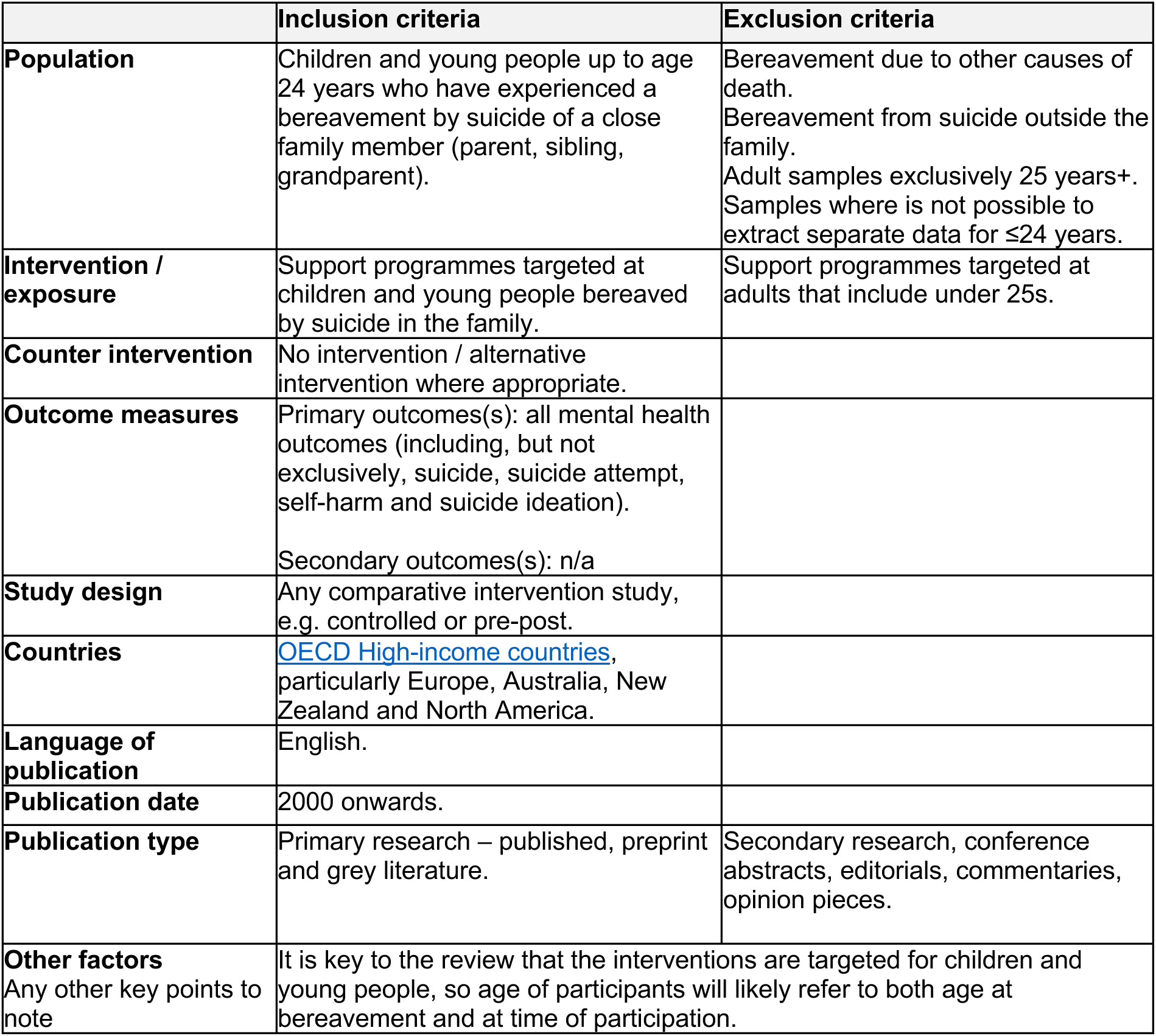
Inclusion and exclusion criteria.

|  | Inclusion criteria | Exclusion criteria |
| --- | --- | --- |
| **Population** | Children and young people up to age 24 years who have experienced a bereavement by suicide of a close family member (parent, sibling, grandparent). | Bereavement due to other causes of death.<br>Bereavement from suicide outside the family.<br>Adult samples exclusively 25 years+.<br>Samples where it is not possible to extract separate data for $\leq 24$ years. |
| **Intervention / exposure** | Support programmes targeted at children and young people bereaved by suicide in the family. | Support programmes targeted at adults that include under 25s. |
| **Counter intervention** | No intervention / alternative intervention where appropriate. |  |
| **Outcome measures** | Primary outcomes(s): all mental health outcomes (including, but not exclusively, suicide, suicide attempt, self-harm and suicide ideation).<br>Secondary outcomes(s): n/a |  |
| **Study design** | Any comparative intervention study, e.g. controlled or pre-post. |  |
| **Countries** | OECD High-income countries, particularly Europe, Australia, New Zealand and North America. |  |
| **Language of publication** | English. |  |
| **Publication date** | 2000 onwards. |  |
| **Publication type** | Primary research – published, preprint and grey literature. | Secondary research, conference abstracts, editorials, commentaries, opinion pieces. |
| **Other factors**<br>Any other key points to note | It is key to the review that the interventions are targeted for children and young people, so age of participants will likely refer to both age at bereavement and at time of participation. |  |

### 5.2 Literature search

Prior to planning this review, a preliminary review of existing reviews was conducted. The findings were presented to the stakeholders and used to refine the scope of the present rapid review of primary studies, and to inform the methods. For details of all the resources searched, please refer to Appendix 1.

A comprehensive search was designed in Medline (see Appendix 2) to identify relevant primary studies and was then translated to the databases listed in Appendix 1. This used a combination of text words, thesaurus terms and medical subject headings. Known literature included relevant studies extracted from the reviews identified during the preliminary searches for existing reviews, and any reports highlighted by the stakeholders.

The grey literature search consisted of reports identified by the review team or provided by stakeholders. Additionally, a search of grey literature websites was generated in collaboration with the stakeholders. For searching grey literature resources, a broad search using word variations of the terms: ‘suicide’, ‘bereavement’, ‘child suicide’, ‘child bereavement’, ‘young people suicide’, ‘young people bereavement’, ‘taken own life’ was conducted.

Searching was completed on the 5th of April 2023.Database searches were imported into EndNote 20 and deduplicated by a single reviewer. Grey literature search results were added to an EndNote library and cross-checked against the database master EndNote library.

### 5.3 Study selection process

The final deduplicated EndNote library was imported into Rayyan, and screening was conducted by two independent review authors. Eligibility criteria was used to assess the titles and abstracts and then full text of all sources identified by the search. The full-text study selection was conducted independently by two reviewers. Grey literature reports were also assessed for eligibility by an individual reviewer and were checked by a second reviewer. The reference lists of any identified systematic reviews were also scanned for any additional relevant primary research.

### 5.4 Data extraction

From each study, the following information was extracted: author(s), year, country, study design, study aim, aim of intervention, type of intervention, data collection methods, sample size, participants, inclusion criteria, setting, geographic location, dates of data collection, key findings. One reviewer carried out data extraction and another reviewer checked the accuracy. Please see Section 6.2 for more information.

### 5.5 Study design classification

This review included all comparative intervention studies located during the systematic search, e.g. controlled or pre-post.

### 5.6 Quality appraisal

The methodological quality of included studies was assessed for the trustworthiness, relevance and results reported using the following critical appraisal tools:

- JBI Critical Appraisal Checklist for quasi-experimental studies
- NHLBI Quality Assessment Tool for Before-After (Pre-Post) Studies With No Control Group

One reviewer assessed the methodological quality of the included studies and a second reviewer verified the judgement. Any disagreements were resolved through discussion. All quality assessment data are presented in the critical appraisal data tables (Tables 5/6, Section 6.3).

### 5.7 Synthesis

A narrative approach was used, including tables detailing the extracted data, to provide descriptive summaries of the selected studies to the reader. This type of analysis is recommended for rapid reviews (Grant and Booth 2009).

### 5.8 Assessment of body of evidence

Due to the paucity of evidence, we were unable to undertake any assessment of the outcomes using GRADE.

## 6. EVIDENCE

### 6.1 Search results and study selection

**Figure 1.**
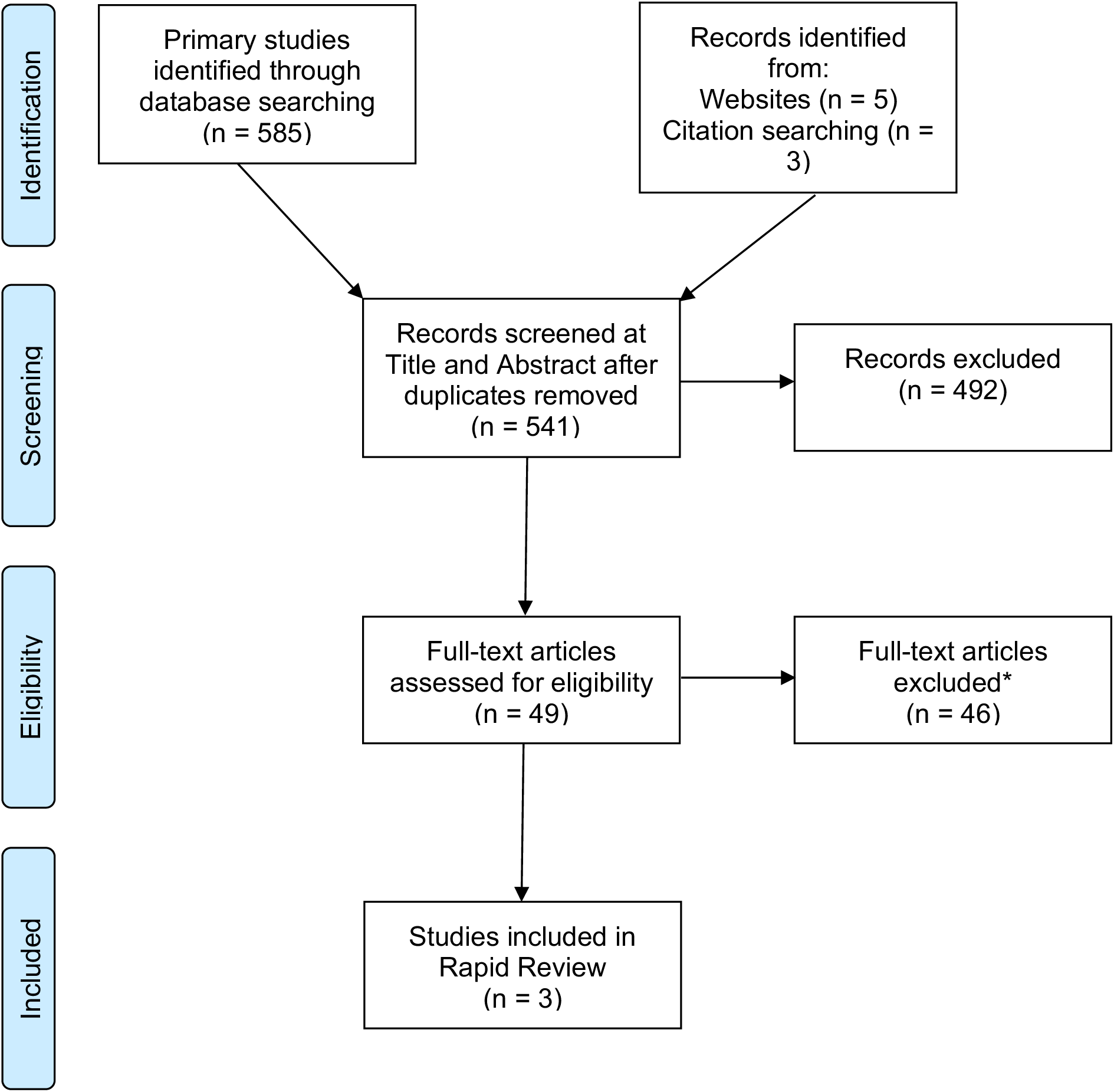
The PRISMA flow diagram. *\*See tables of excluded studies in appendix 4*

### 6.2 Data extraction

**Table 4.**
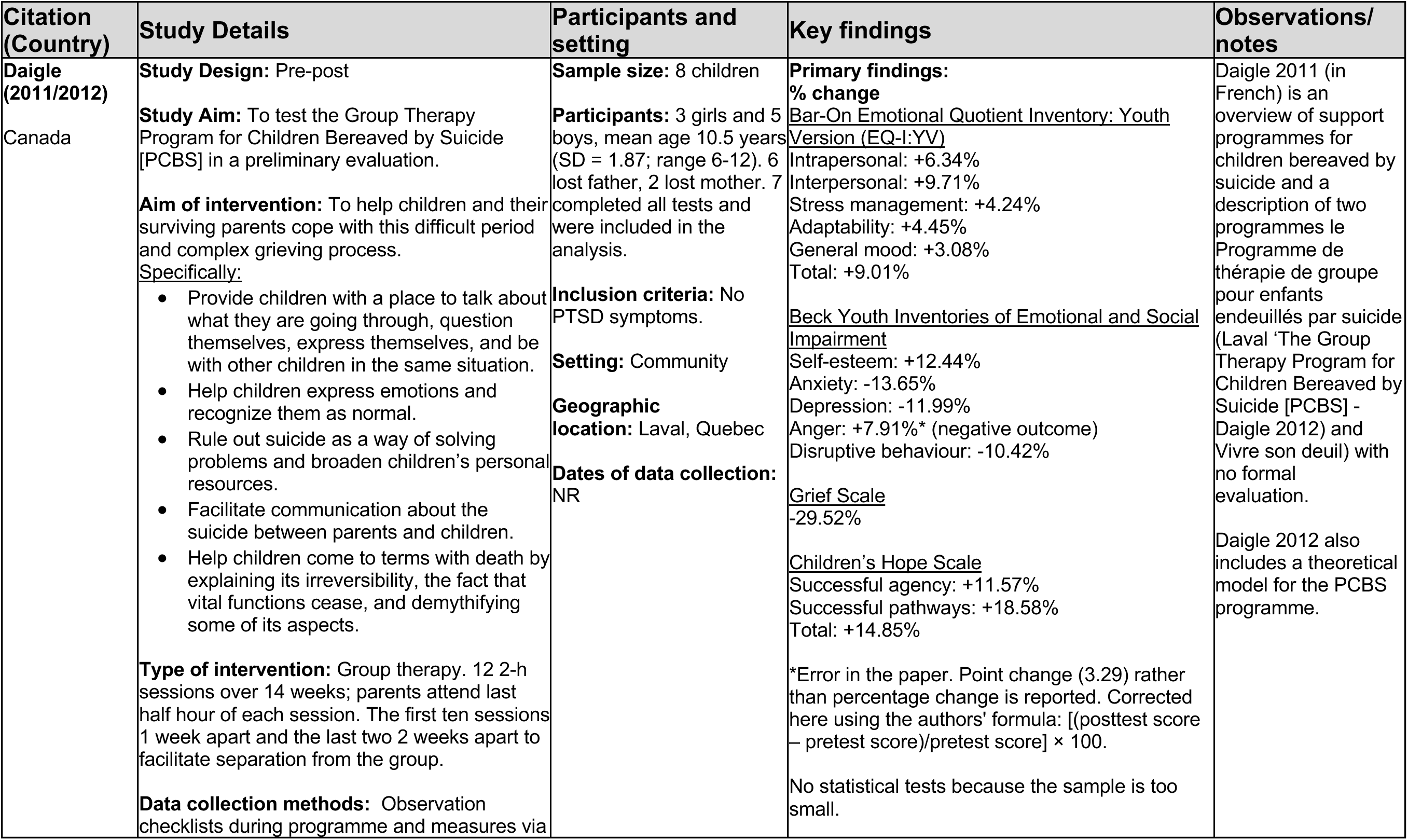
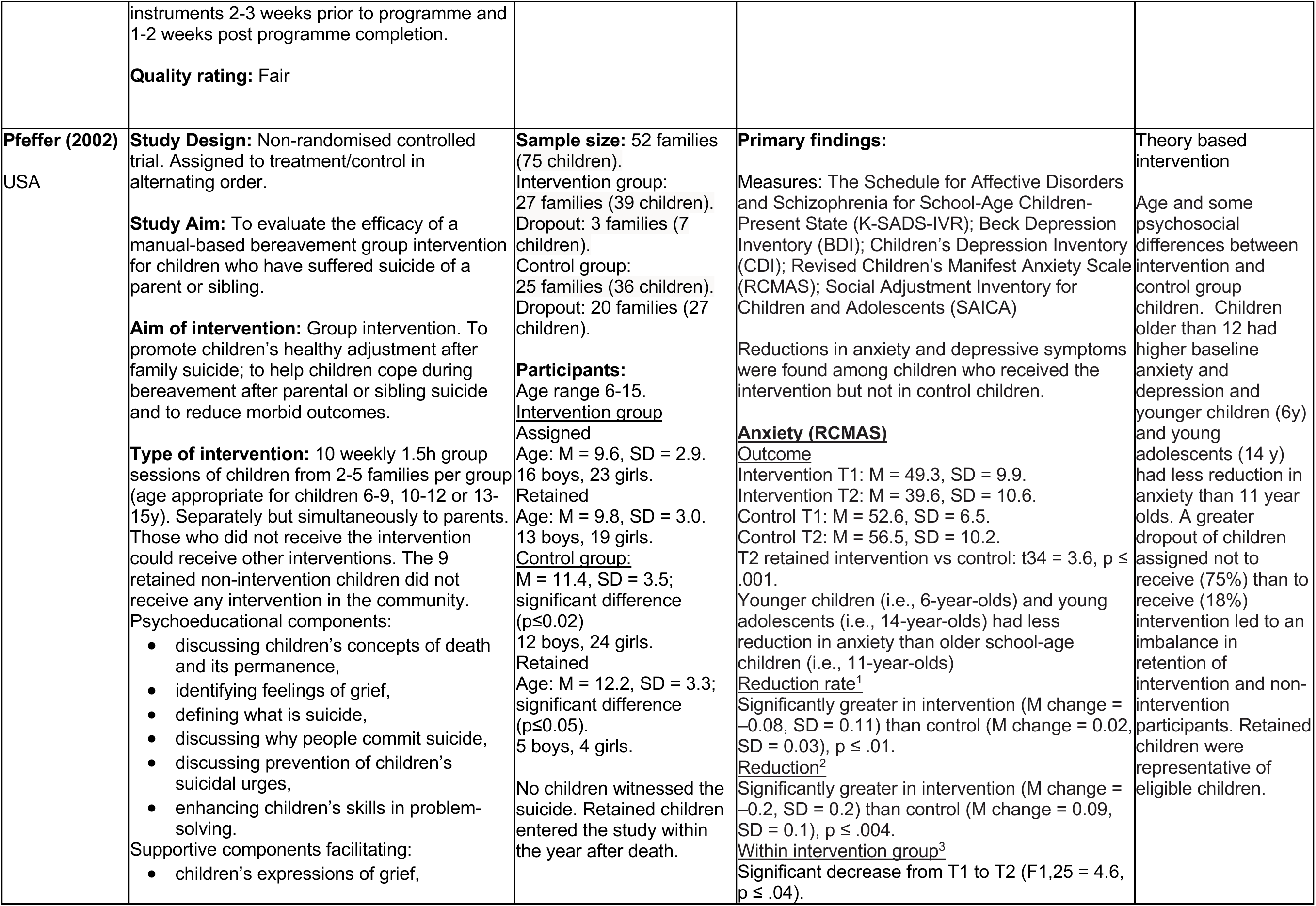
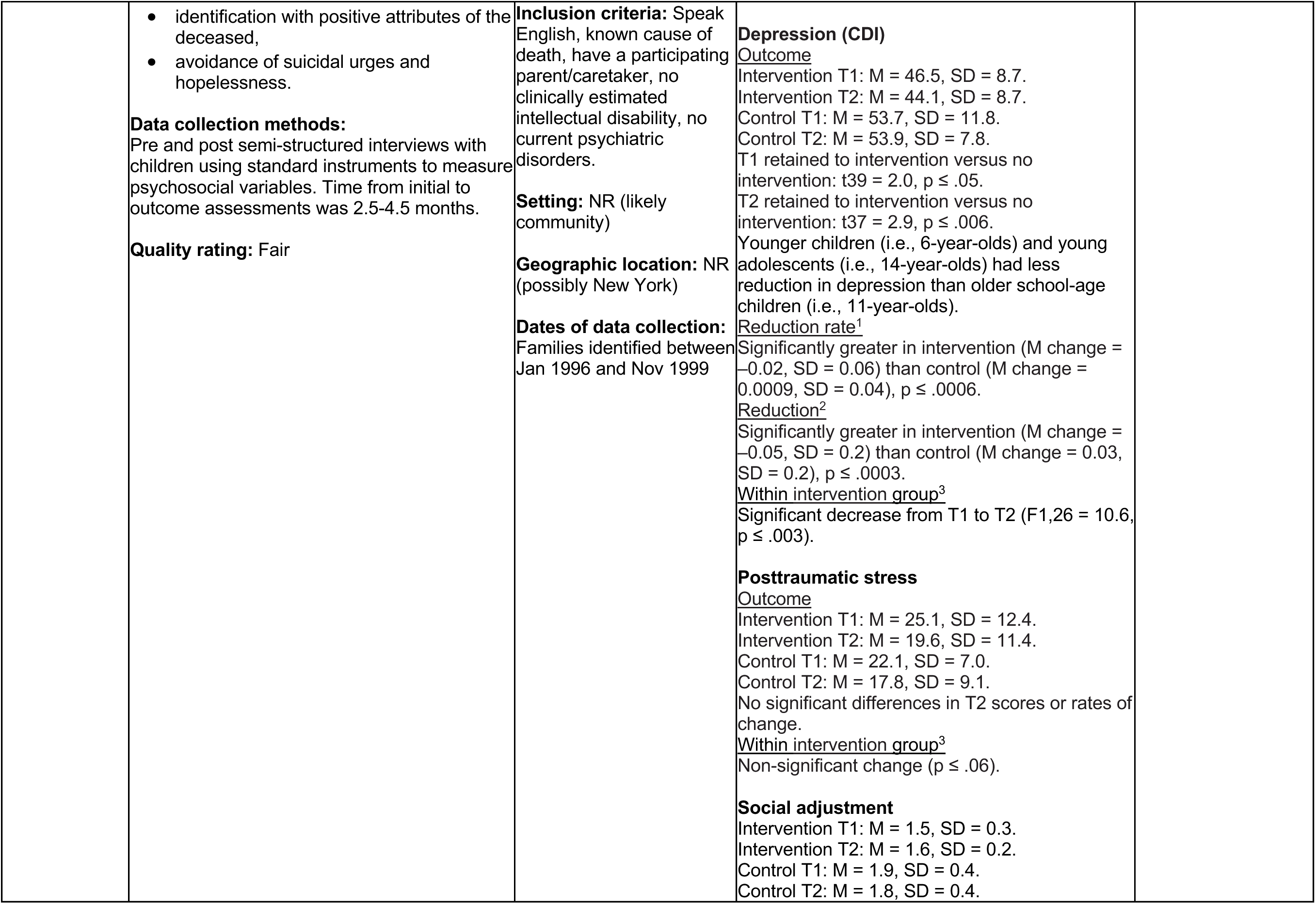
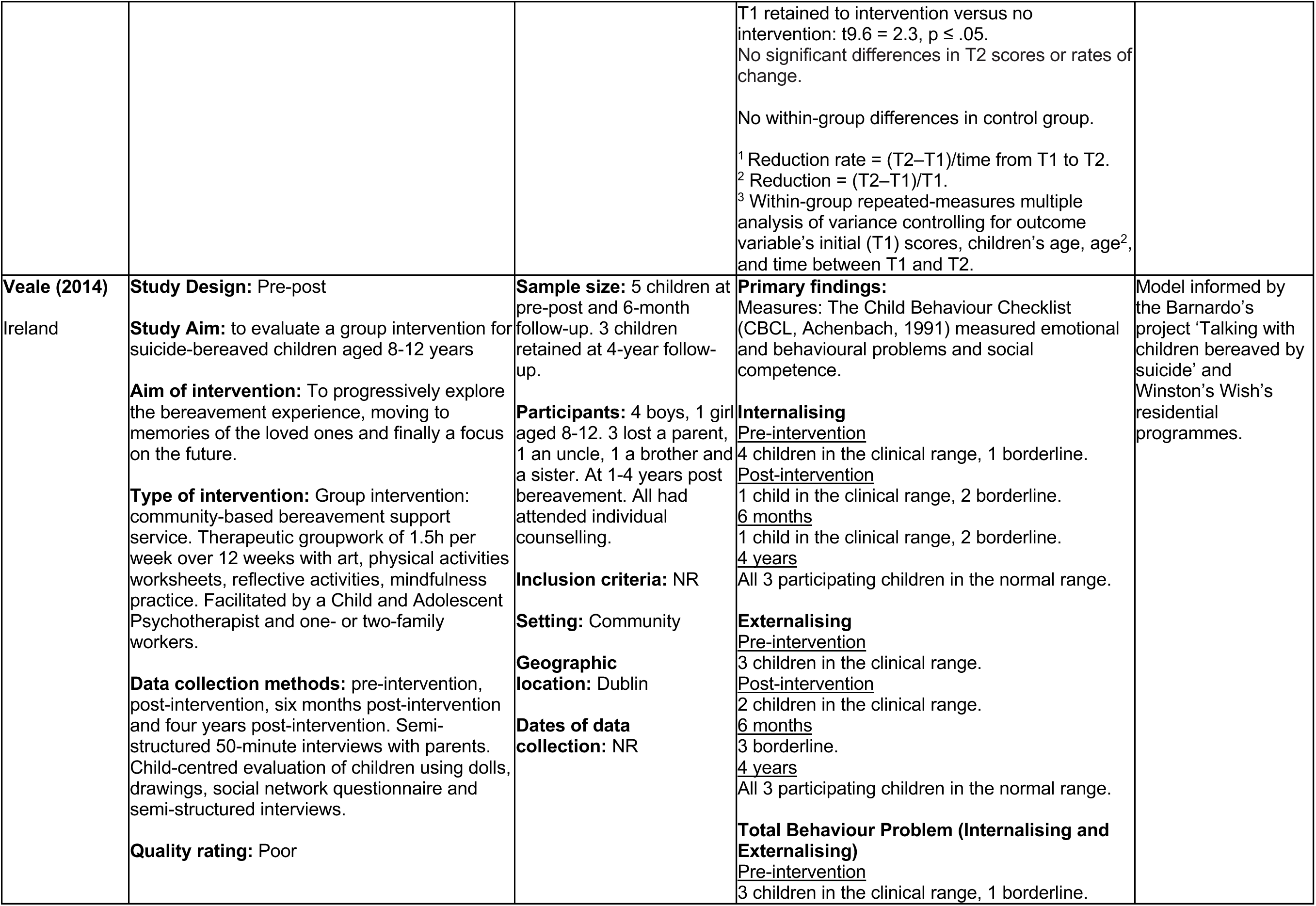
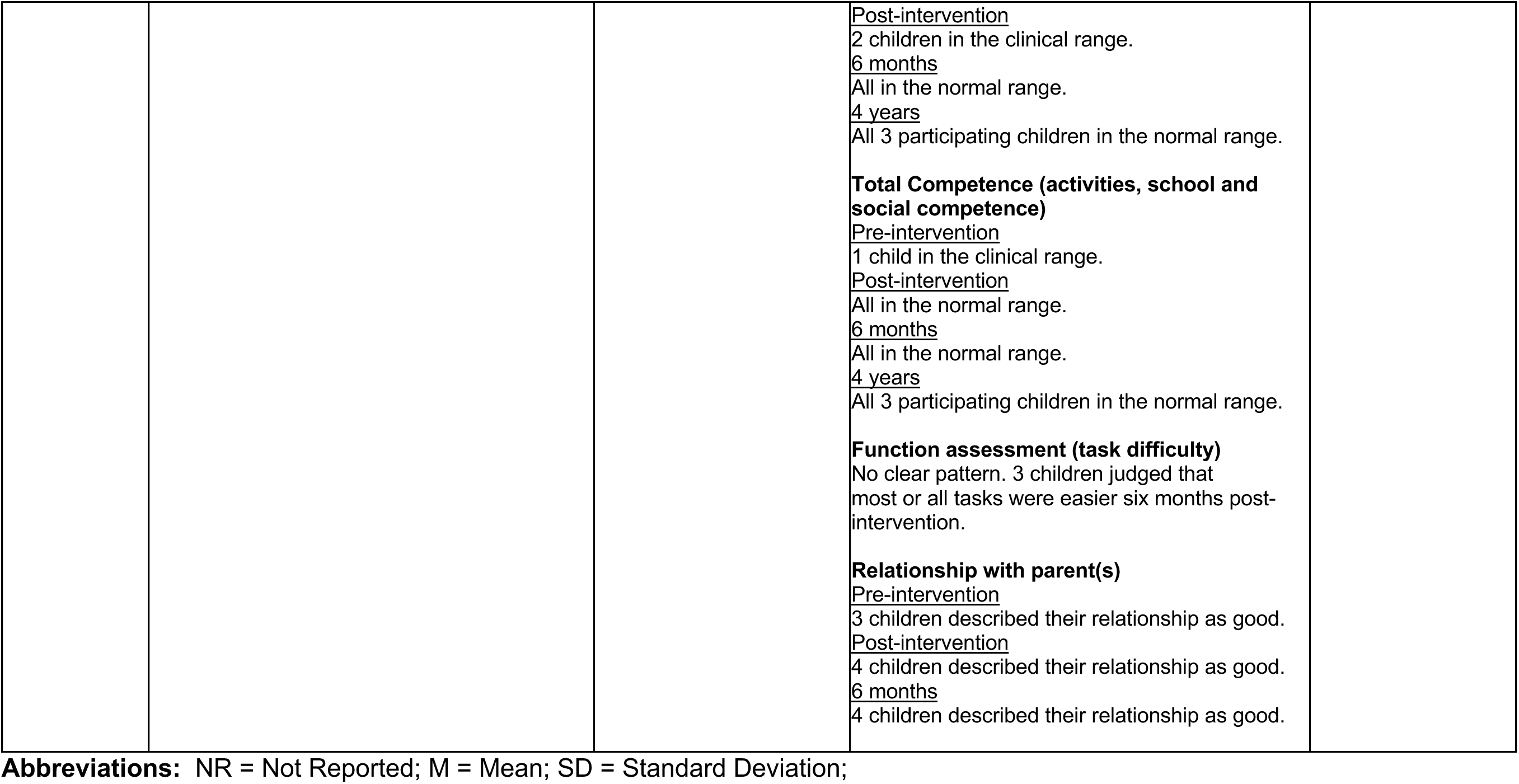
Data extracted from included studies.

### 6.3 Quality appraisal

**Table 5.** Critical appraisal of Pre-Post intervention studies (NHIBI Checklist for Before-After (pre-post) studies with no control group).

| Questions | Daigle 2012 | Veale 2014 |
| --- | --- | --- |
| 1. Was the study question or objective clearly stated? | Yes | Yes |
| 2. Were eligibility/selection criteria for the study population prespecified and clearly described? | Yes | No |
| 3. Were the participants in the study representative of those who would be eligible for the test/service/intervention in the general or clinical population of interest? | Yes | Yes |
| 4. Were all eligible participants that met the prespecified entry criteria enrolled? | No | NR |
| 5. Was the sample size sufficiently large to provide confidence in the findings? | No | No |
| 6. Was the test/service/intervention clearly described and delivered consistently across the study population? | Yes | Yes |
| 7. Were the outcome measures prespecified, clearly defined, valid, reliable, and assessed consistently across all study participants? | Yes | Yes |
| 8. Were the people assessing the outcomes blinded to the participants' exposures/interventions? | No | No |
| 9. Was the loss to follow-up after baseline 20% or less? Were those lost to follow-up accounted for in the analysis? | No | Yes |
| 10. Did the statistical methods examine changes in outcome measures from before to after the intervention? Were statistical tests done that provided p values for the pre-to-post changes? | No | No |
| 11. Were outcome measures of interest taken multiple times before the intervention and multiple times after the intervention (i.e., did they use an interrupted time-series design)? | No | No |
| 12. If the intervention was conducted at a group level (e.g., a whole hospital, a community, etc.) did the statistical analysis take into account the use of individual-level data to determine effects at the group level? | Yes | No |
| **Overall appraisal comments** | Quality: Fair | Quality: Poor |

**Table 6.** Critical appraisal of Non-randomised controlled trials (JBI Checklist for quasi-experimental studies).

| Questions | Pfeffer 2002 |
| --- | --- |
| 1. Is it clear in the study what is the 'cause' and what is the 'effect' (i.e. there is no confusion about which variable comes first)? | Yes |
| 2. Were the participants included in any comparisons similar? | No |
| 3. Were the participants included in any comparisons receiving similar treatment/care, other than the exposure or intervention of interest? | Yes |
| 4. Was there a control group? | Yes |
| 5. Were there multiple measurements of the outcome both pre and post the intervention/exposure? | No |
| 6. Was follow up complete and if not, were differences between groups in terms of their follow up adequately described and analyzed? | Yes |
| 7. Were the outcomes of participants included in any comparisons measured in the same way? | Yes |
| 8. Were outcomes measured in a reliable way? | Yes |
| 9. Was appropriate statistical analysis used? | Yes |
| **Overall appraisal comments** | Quality: Fair |

### 6.4 Information available on request

The protocol, all search strategies, details of excluded studies and individual critical appraisal forms are available from.

## 7.2 Acknowledgements

The Health and Care Research Wales Evidence Centre and SURE team would like to thank stakeholders for their involvement, time and expertise in this rapid review process: Claire Cotter, Ann John and Rashmi Kumar.

## 8. APPENDIX

### 8.1 APPENDIX 1: Resources searched during Rapid Review Searching

| Resource | Number of hits |
| --- | --- |
| **Database resources searched** |  |
| OVID - Medline | 118 |
| OVID - PsycInfo | 165 |
| ProQuest Social Care<br>ProQuest - Sociology Collection:<br>1. Applied Social Sciences Index and Abstracts (ASSIA)<br>2. Sociological Abstracts<br>3. Social Services Abstracts<br>4. Sociology Database | 42 |
| ASSIA via ProQuest | 61 |
| Cinahl via EBSCO Host | 140 |
| Scopus via Elsevier | 242 |
| Total | 768 |
| **Total after Deduplication** | **531** |
| **Grey literature websites** |  |
| <u>2 Wish</u> | 0 |
| <u>Anna Freud National Centre for Children and Families</u> | 0 |
| <u>Barnardo's</u> | 1 |
| <u>Child Bereavement UK</u> | 0 |
| <u>Childhood bereavement Network</u> | 1 |
| <u>Childline</u> | 0 |
| <u>Children and Young People's Commissioner (Scotland)</u> | 0 |
| <u>Children's Commissioner (England)</u> | 0 |
| <u>Children's Commissioner (Wales)</u> | 0 |
| <u>Cruse</u> | 0 |
| <u>Grief Encounter</u> | 0 |
| <u>Hope Again (youth site of Cruse)</u> | 0 |
| <u>If U Care Share</u> | 0 |
| <u>Jigsaw 4 U</u> | 0 |
| <u>Mind</u> | 0 |
| <u>National Suicide Prevention Alliance</u> | 1 |
| <u>National Youth Agency</u> | 0 |
| <u>NICCY – Commissioner for Children and Young People (Northern Ireland)</u> | 0 |
| <u>NICE</u> | 0 |
| <u>NSPCC</u> | 0 |
| <u>PAPYRUS UK</u> | 0 |
| <u>Royal College of Psychiatrists</u> | 0 |
| <u>Samaritans</u> | 0 |
| <u>Suicide bereavement UK</u> | 0 |
| <u>Support After Suicide Partnership</u> | 0 |
| <u>Survivors of Bereavement by Suicide</u> | 1 |
| <u>The Children's society</u> | 0 |
| <u>Trauma Council</u> | 0 |
| <u>UK Government</u> | 0 |
| <u>Welsh Government</u> | 1 |
| <u>Winston's Wish</u> | 0 |

### 8.2 APPENDIX 2: Search strategy (OVID Medline)

Ovid MEDLINE(R) ALL <1946 to March 27, 2023>

1. Suicide/ or suicide, completed/ 40095
2. ((take or took or taken) adj2 own life).tw. 62
3. ((self-injur* adj3 death) or (self-injur* adj3 suicide attempt)).mp. [mp=title, book title, abstract, original title, name of substance word, subject heading word, floating sub-heading word, keyword heading word, organism supplementary concept word, protocol supplementary concept word, rare disease supplementary concept word, unique identifier, synonyms, population supplementary concept word, anatomy supplementary concept word] 93
4. or/1-3 40210
5. (parent* or mother* or mum* or mom* or dad* or father* or caregiver* or grandparent* or grandmother* or grandfather* or grandpa or grandma).tw. 1183327
6. (family adj2 member*).tw. 113410
7. (sibling* or sister* or brother*).tw. 106848
8. 5 or 6 or 7 1355644
9. 4 and 8 1942
10. exp Child/ or (child* or adolescen* or teenage* or (young adj person*) or (young adj people) or youth or (young adj adult*) or student*).tw. 3216990
11. (boy* or girl* or kid* or preteen* or pre teen* or preadolesc* or pre adolesc* or juvenil* or schoolchild* or teen* or youth or young adolesc* or (young adj boy) or (young adj man) or (young adj girl) or (young adj woman)).tw. 1036250
12. exp Young Adult/ 1006685
13. 10 or 11 or 12 4555515
14. Bereavement/ 6588
15. (grief or griev* or bereav*).tw.17193
16. mourn*.tw. 2157
17. or/14-16 20148
18. 9 and 13 and 17 160
19. (comment or editorial or letter).pt. 2145324
20. 18 not 19 159
21. (meta-analysis or meta analysis or review or systematic review).pt.3289088
22. 20 not 21 143
23. limit 22 to (english language and yr=“2000 –Current”) 118

### 8.3 APPENDIX 3: Data extraction for additional studies outside the inclusion criteria

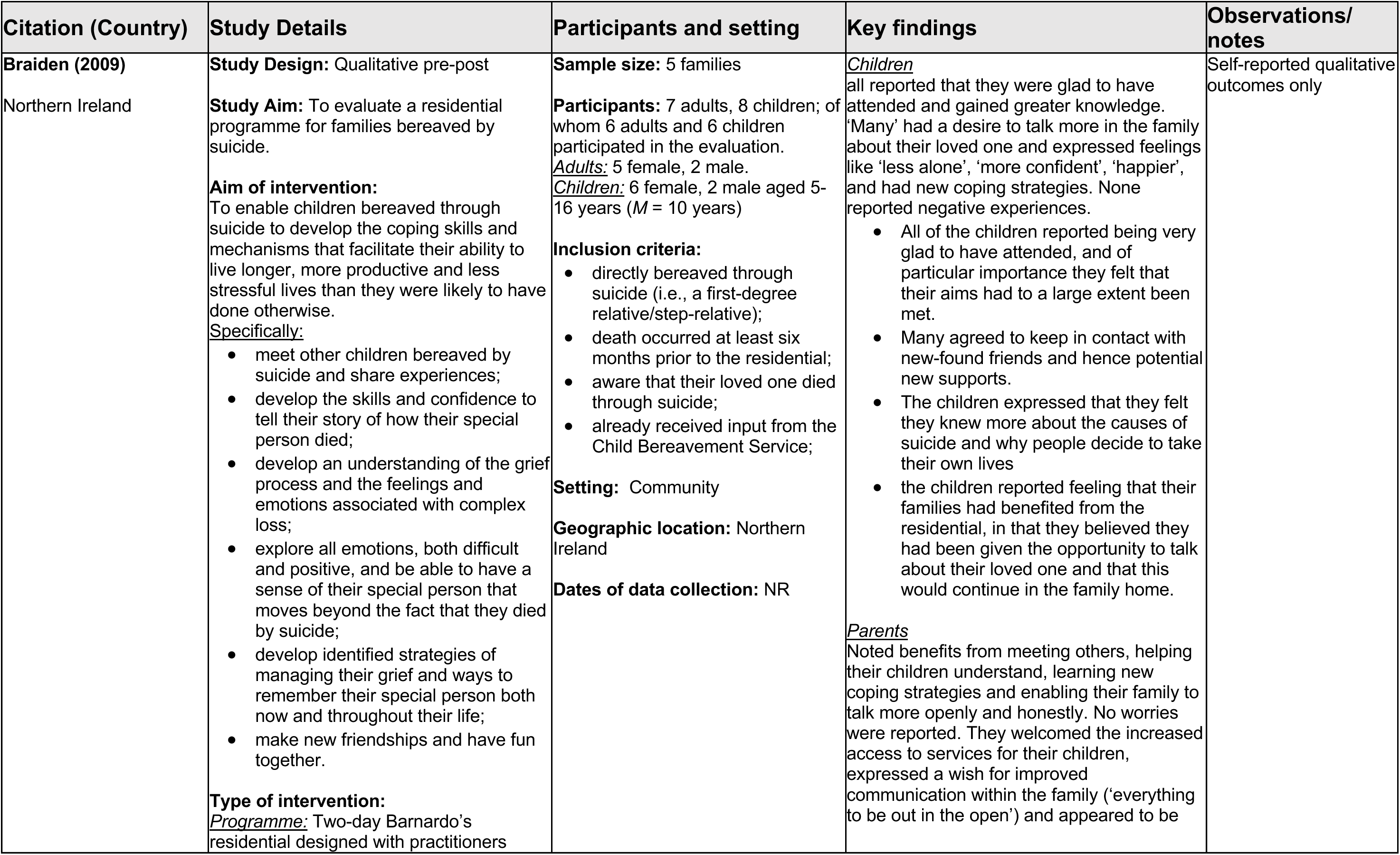
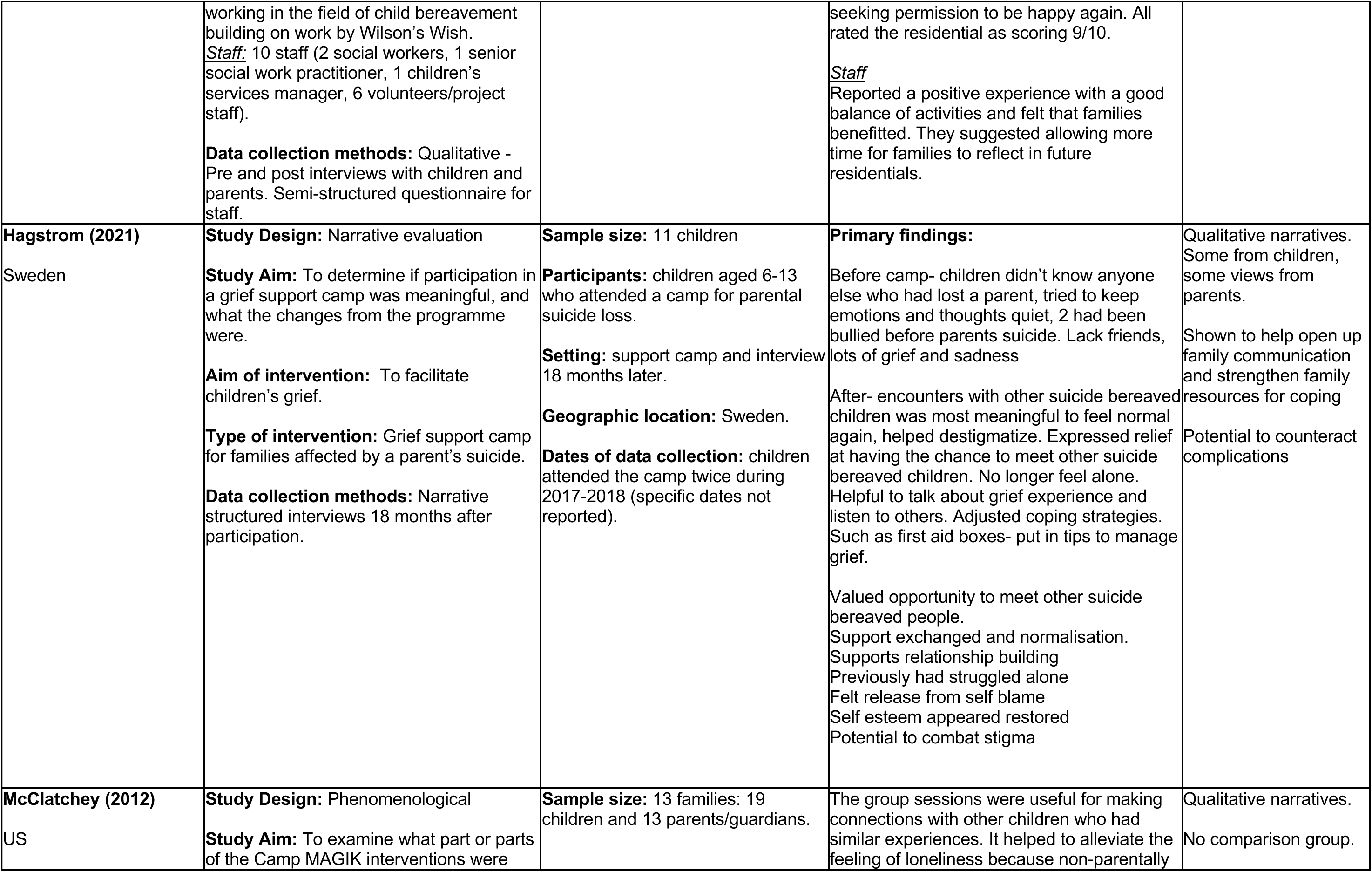
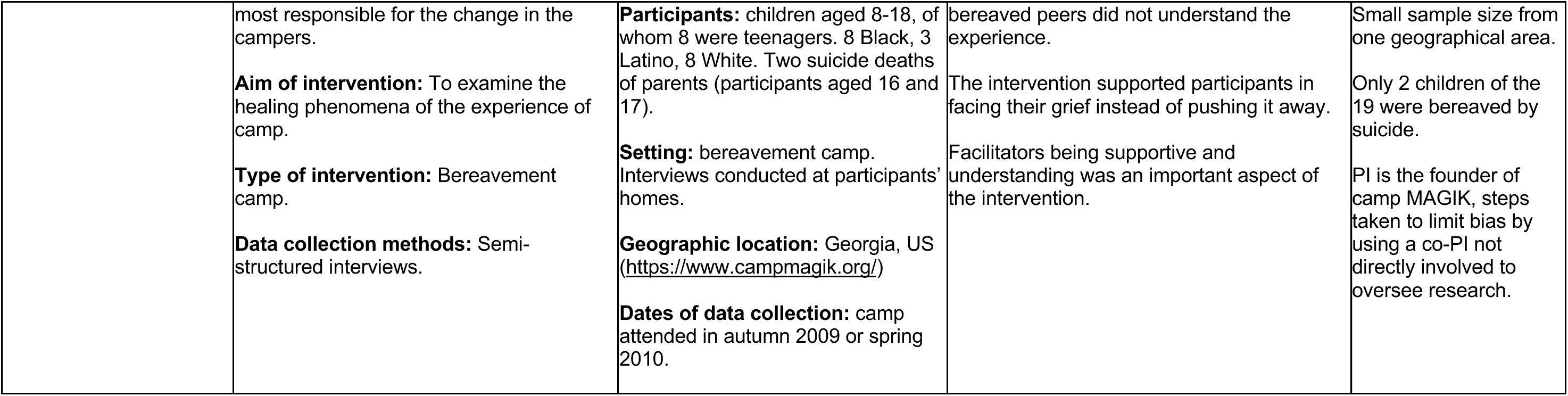

### 8.4 APPENDIX 4: Full list of excluded studies

**Table 7.**
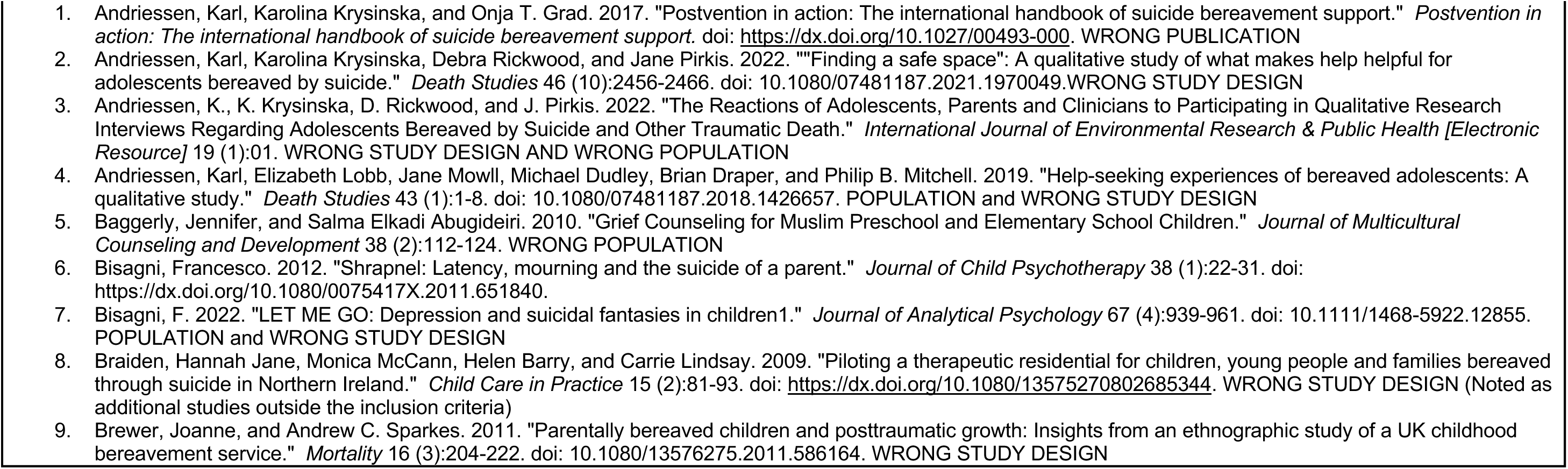
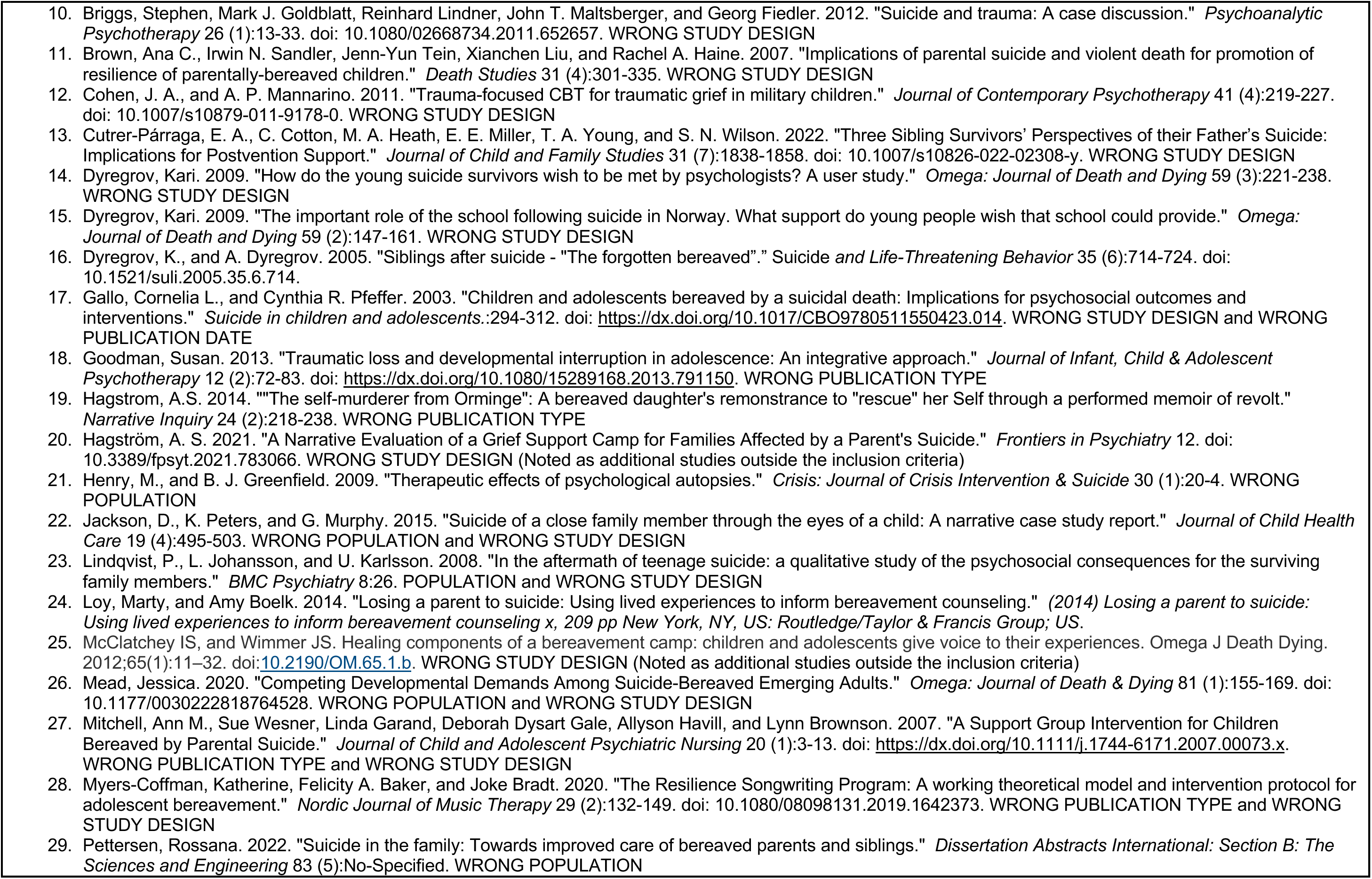
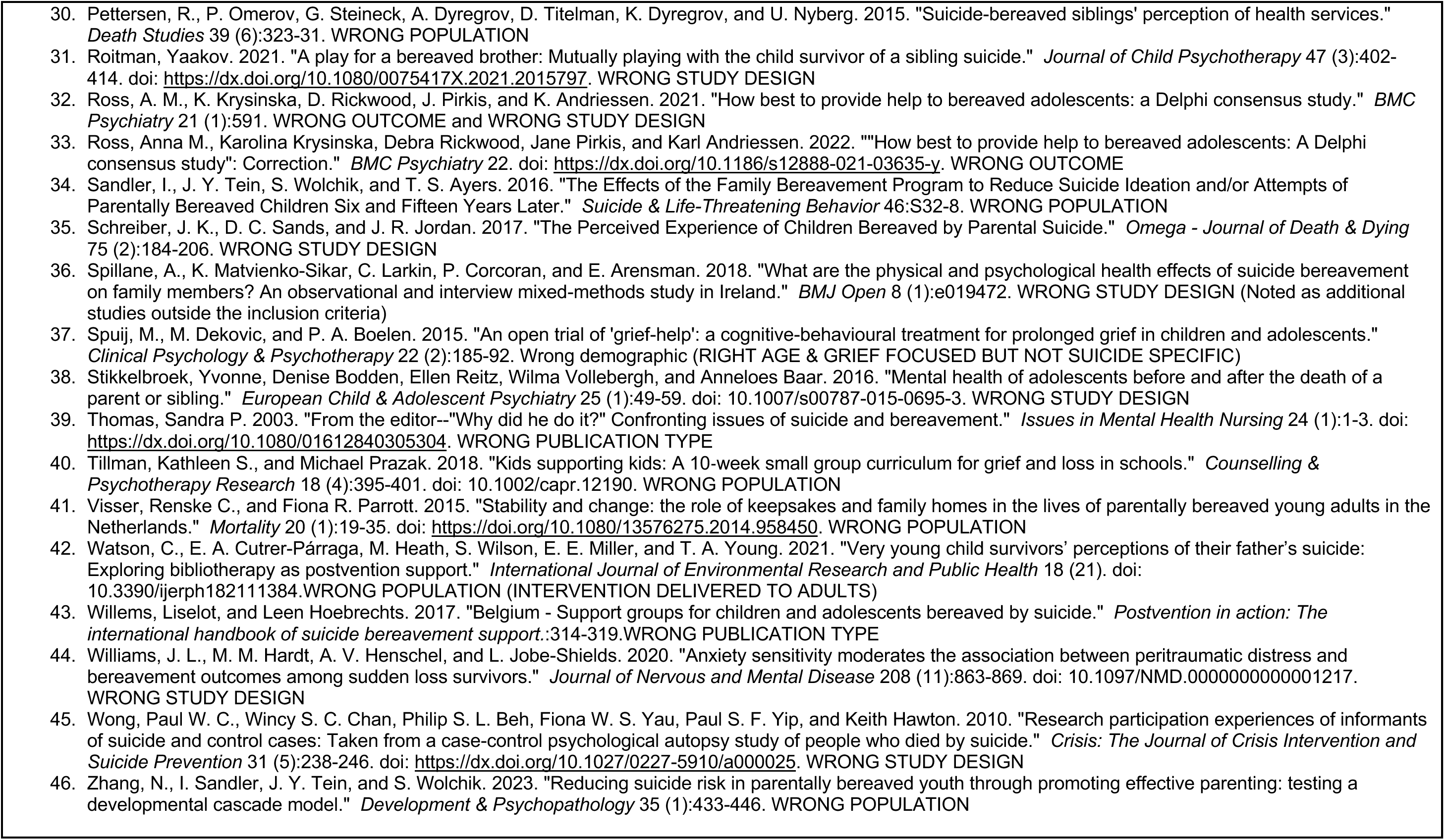
Excluded studies from database searches.

**Table 8.** Excluded studies from grey literature searches.

|  |  |
| --- | --- |
| Barnardo's | An overview of evidence-based interventions – For children and young people experiencing bereavement, loss and grief.<br>https://www.barnardos.org.uk/sites/default/files/2021-02/MHF%20Scotland%20Mapping%20interventions%20bereavment.pdf - Wrong population (not suicide specific) |
| National Suicide Prevention Alliance | Suicide bereavement in the UK: Descriptive findings from a national survey<br>https://nspa.org.uk/wp-content/uploads/2022/05/Suicide-bereavement-in-the-UK.pdf - wrong population |
| Welsh Government | A Scoping Survey of Bereavement Services in Wales end of study report<br>https://www.gov.wales/sites/default/files/publications/2019-12/scoping-survey-of-bereavement-services-in-wales-report.pdf?_ga=2.44440038.39583088.1679048462-5037011.1679048462 - wrong population |

**Table 9.**
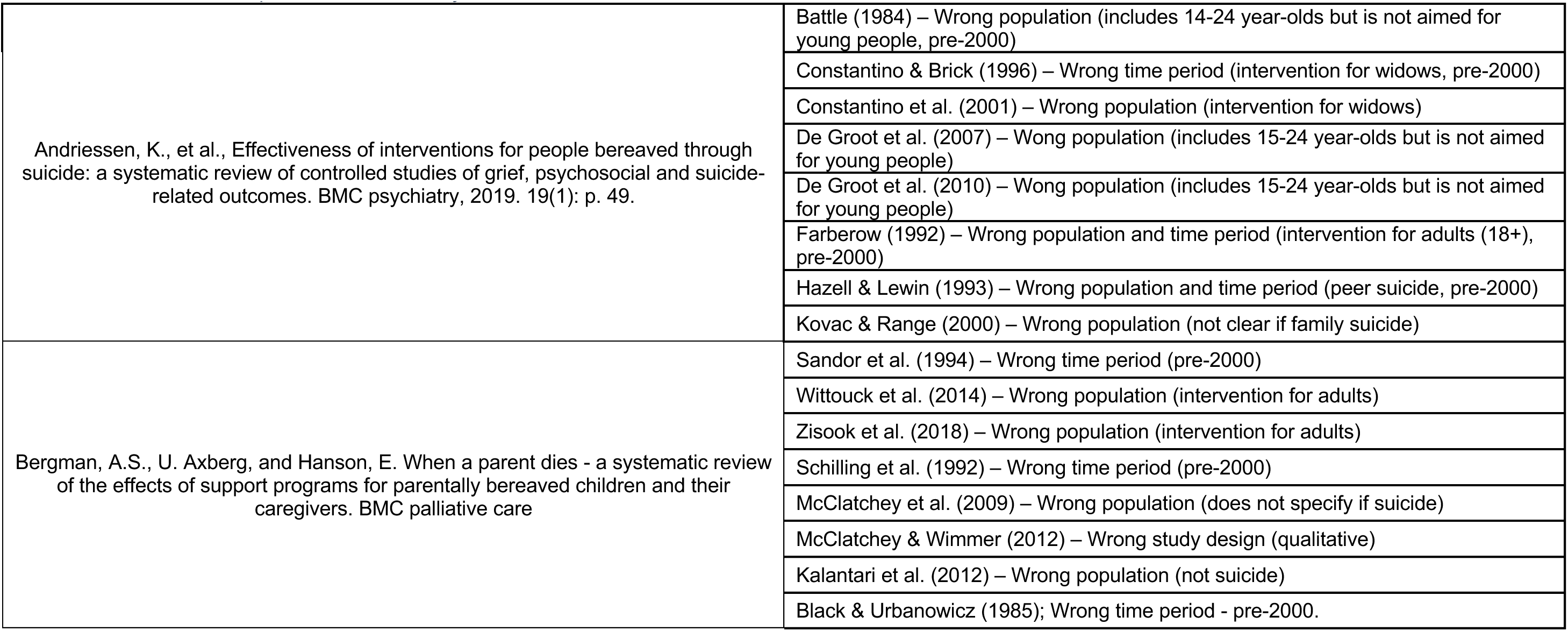
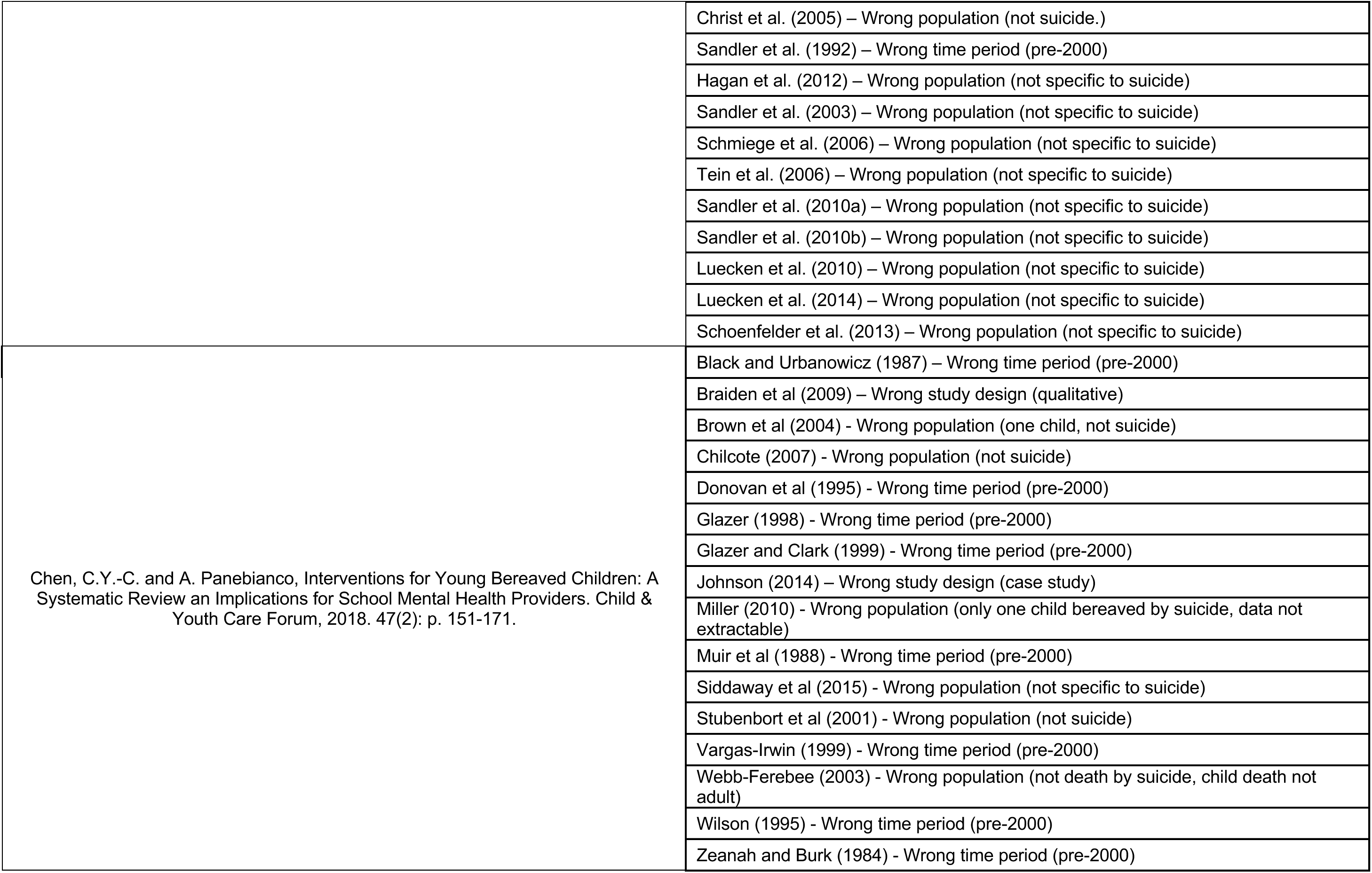
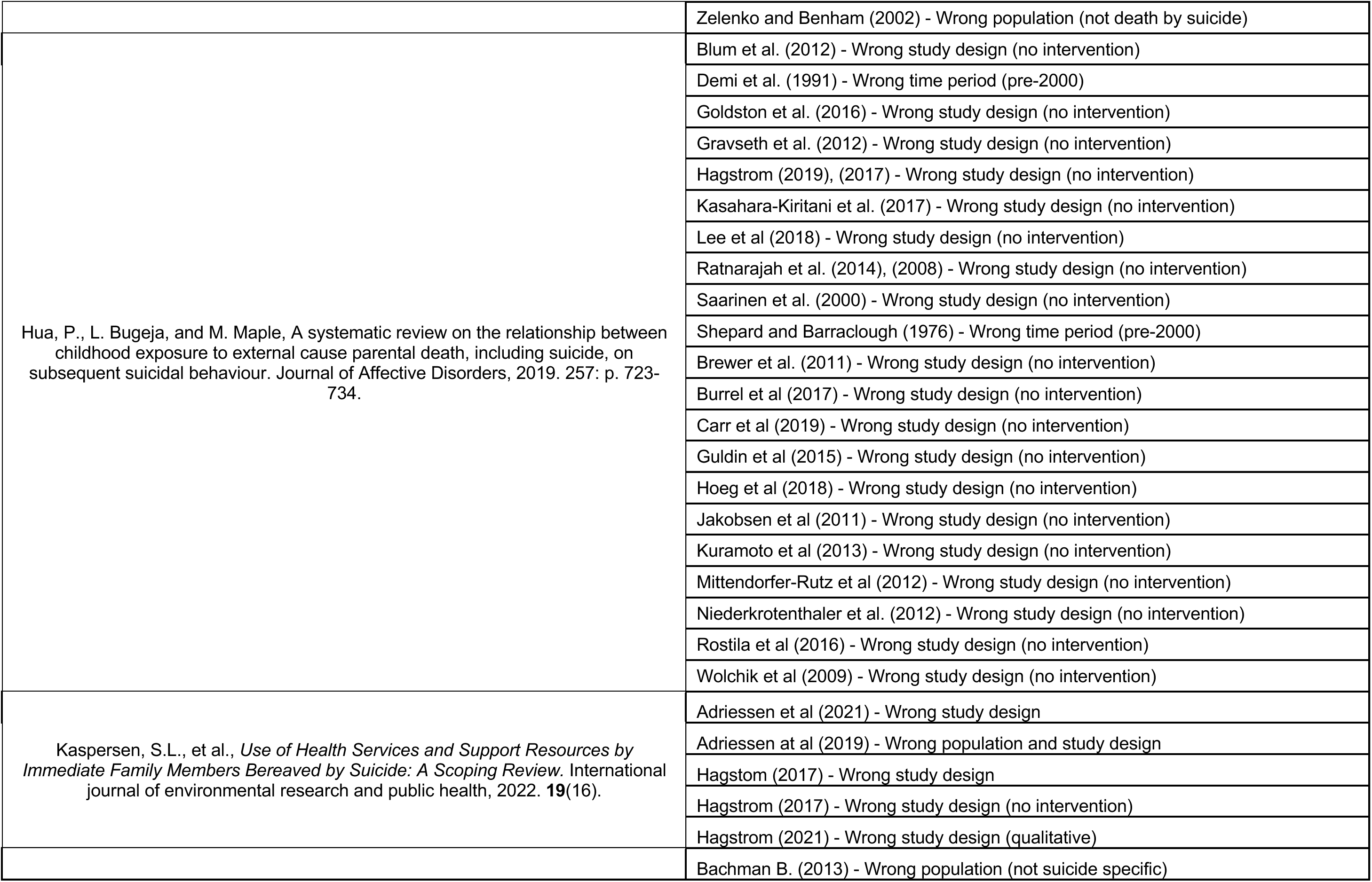
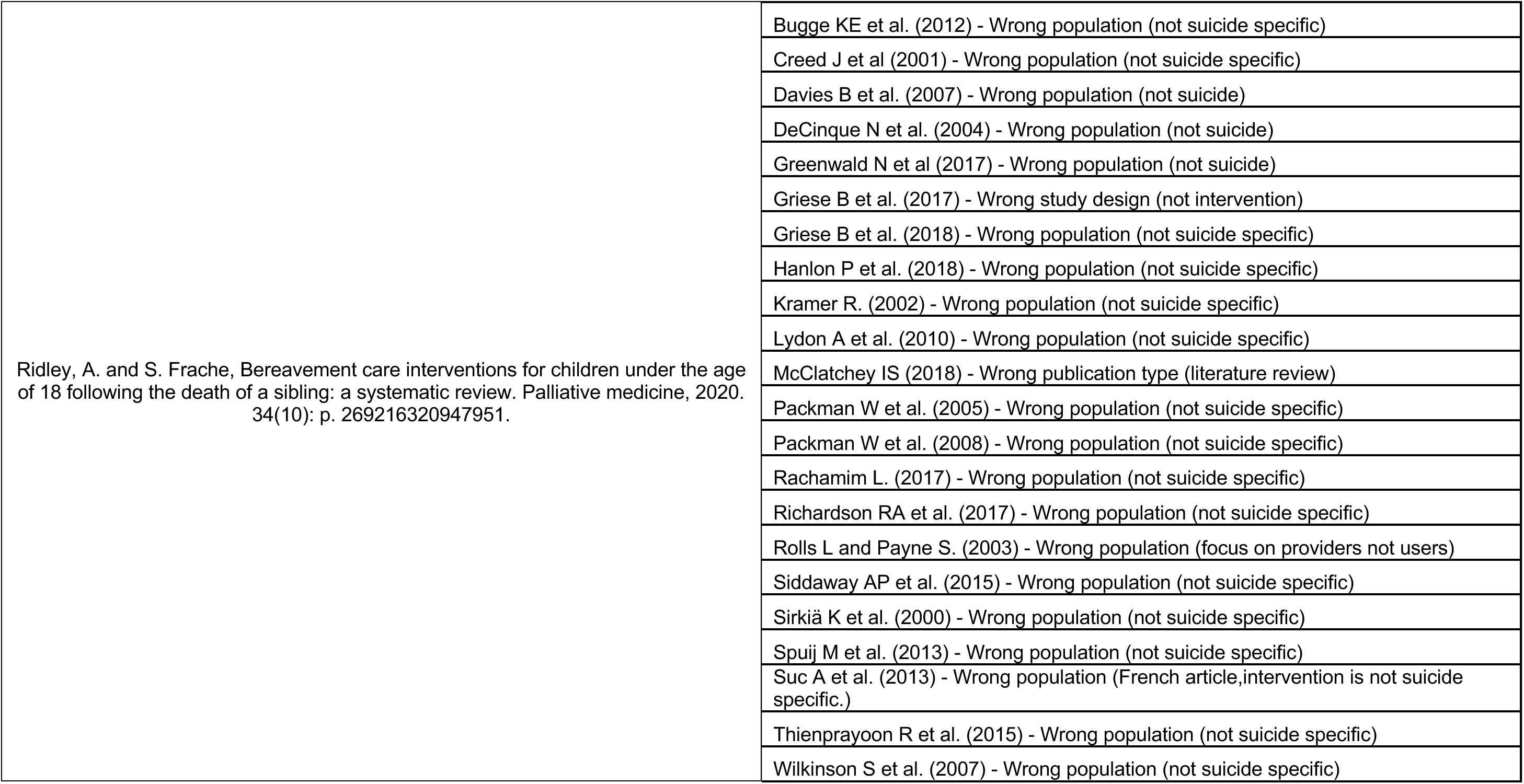
Excluded studies unpicked from secondary research.

